# Multimodal mental health assessment with remote interviews using facial, vocal, linguistic, and cardiovascular patterns

**DOI:** 10.1101/2023.09.11.23295212

**Authors:** Zifan Jiang, Salman Seyedi, Emily Griner, Ahmed Abbasi, Ali Bahrami Rad, Hyeokhyen Kwon, Robert O. Cotes, Gari D. Clifford

## Abstract

**Objective:** The current clinical practice of psychiatric evaluation suffers from subjectivity and bias, and requires highly skilled professionals that are often unavailable or unaffordable. Objective digital biomarkers have shown the potential to address these issues. In this work, we investigated whether behavioral and physiological signals, extracted from remote interviews, provided complimentary information for assessing psychiatric disorders.

**Methods:** Time series of multimodal features were derived from four conceptual modes: facial expression, vocal expression, linguistic expression, and cardiovascular modulation. The features were extracted from simultaneously recorded audio and video of remote interviews using task-specific and foundation models. Averages, standard deviations, and hidden Markov model-derived statistics of these features were computed from 73 subjects. Four binary classification tasks were defined: detecting 1) any clinically-diagnosed psychiatric disorder, 2) major depressive disorder, 3) self-rated depression, and 4) self-rated anxiety. Each modality was evaluated individually and in combination.

**Results:** Statistically significant feature differences were found between controls and subjects with mental health conditions. Correlations were found between features and self-rated depression and anxiety scores. Visual heart rate dynamics achieved the best unimodal performance with areas under the receiver-operator curve (AUROCs) of 0.68-0.75 (depending on the classification task). Combining multiple modalities achieved AUROCs of 0.72-0.82. Features from task-specific models outperformed features from foundation models.

**Conclusion:** Multimodal features extracted from remote interviews revealed informative characteristics of clinically diagnosed and self-rated mental health status.

**Significance:** The proposed multimodal approach has the potential to facilitate objective, remote, and low-cost assessment for low-burden automated mental health services.

## I. Introduction

The World Health Organization estimated in 2019 that 13% of the world population, or close to one billion people worldwide, live with a mental disorder, where most of them do not have access to effective care [1]. Since the COVID-19 pandemic, those numbers have been rising rapidly, and the pandemic continued to impede access to already underserved psychiatric health services [2]–[4]. In addition to being the second most common cause of years of life lived with disability worldwide [5], this crisis of psychiatric disorders translates to an economic burden of $280 billion every year in the United States alone [6]. To reduce the high yearly cost and to delay the transition into often chronic or life-long psychiatric conditions, it is critical to gain a better understanding and to provide objective, fast, and accessible detection of those disorders to enable early and effective interventions. However, the present diagnosis and phenotyping of psychiatric disorders fail to fully satisfy this dire need due to its subjectivity and biases, and access to psychiatric care is limited even in high-income countries such as the US [7].

Currently, psychiatric disorders such as depression and anxiety disorder are diagnosed through the subjective clinical evaluation of signs and symptoms established by the Diagnostic and Statistical Manual of Mental Disorders (DSM-5) [8] or the International Classification of Diseases, 10th revision [9]. These diagnostic criteria often suffer from low inter-rater reliability. In the DSM-5 field trials [10], inter-rater reliability (Cohen’s kappa, *κ*) was just 0.28 for a diagnosis of major depressive disorder (MDD) and 0.20 for general anxiety disorder (GAD). The reliability for a diagnosis of MDD was even lower (*κ* = 0.16) when interviews were carried out by general practitioners [11], who perform large percent of diagnoses worldwide [12] and in the US as well due to the decline of supply of psychiatrists [13]. Factors such as differences in training, biases (race, gender, culture), and interview style were the most common explanations for discrepancies between raters [14], [15]. Self-rated questionnaires such as General Anxiety Disorder-7 (GAD-7) [16], and Patient Health Questionnaire-9 (PHQ-9) [17] are also widely used in practice for initial screening and symptom monitoring purposes. Naturally, these scales are highly subjective as they are self-reported: the symptoms reported tend to be over-reported and more severe than observer ratings and highly depend on the subjective response processes [18].

The rapid development of objective automated digital assessment tools has the potential to aid clinicians in the diagnosis and evaluation of mental illness and to limit the impact of these illnesses on patients and on society [19]. Research groups have developed tools using various types of data modality, validated in numerous mental health populations, including depression [20], [21], anxiety disorder [22], schizophrenia [23], posttraumatic stress disorder (PTSD) [24], and almost all common psychiatric disorders. Diverse modalities of signals have been investigated, including behavioral signals, such as facial and body movements [20], [21], [25], speech acoustics [26]–[28], verbal or written content [29], sleep [30] and activity [24], [31] patterns, as well as physiological signals such as cardiovascular (heart rate [24], [32], electrocardiogram [28], [33], etc.) and neural signals (electroencephalogram [34], [35], functional magnetic resonance imaging [36], [37] and functional near-infrared spectroscopy [38]). The multimodal approach, or the combination of multiple types of signals, has been widely adopted to improve the accuracy and robustness of those automated assessments [39], [40]. For example, [41], [42] combined behavioral signals, including cues from video, audio, and text, while others [24], [43] found the combination and interaction between physiological and behavioral signals were also useful in evaluating disorders.

While the findings in the above studies were promising, there remain unsolved challenges: data in most of them were collected within a lab-controlled environment and/or with specialized hardware, which prohibits potential future access and might not be able to generalize to actual clinical practice. The increasing use of telemedicine in psychiatry in recent years, even further accelerated by the COVID-19 pandemic [44], provided a promising approach to improve the access and effect of psychiatric care [45]–[47], while at the same presented an unprecedented opportunity of data collection for objective psychiatric assessments development without the limitation of geographical location and specialized hardware [48]. This begs the question of whether data collected remotely, such as in [49], [50], and in our previous research protocol [19], can provide a similar level of information as the data collected in a lab-controlled environment.

To address those challenges, we investigated whether each and the combination of behavioral and physiological signals, extracted from audio-visual recordings of remote telehealth interviews, which were collected using heterogeneous generic electronic devices (laptops, tablets, or smartphones) from the patients, were informative in assessing the multiple facets of psychiatric disorders of control subjects and subjects with mental health conditions (MHC).

The main contributions of this work are as follows: (1) We showed that audio-visual recordings of remote interviews collected fully remotely and without device limitation could be used to assess mental health states, with similar performance compared to the performance shown in previous studies where data were collected from lab-controlled environments. (2) We proposed a multimodal machine learning analysis framework, where we extracted both hand-crafted features and self-supervised-learned representations of facial, vocal, linguistic, and remote photoplethysmography (rPPG) patterns using signal processing approaches and state-of-the-art deep learning models, including convolutional neural networks (CNN) and transformer-based [51] foundation models. (3) Using those features and derived temporal dynamics, we compared the performance of features extracted from different modalities, with different models, and the performance of the combined features of multiple modalities, in classifying states of depression, anxiety, and whether mentally healthy (i.e., without any diagnosed disorder) using both self-reported scales (PHQ-9, GAD-7) and diagnoses made by clinicians.

## II. Dataset

### A. Participants

The overall recruitment protocol can be found in Cotes et al. [19], where we planned to recruit three outpatient groups: (1) 50 patients with schizophrenia, (2) 50 patients with unipolar major depressive disorder, and (3) 50 individuals with no psychiatric history. Due to the difficulty of recruiting enough in-person schizophrenia subjects during COVID-19, in this work, we focused on analyzing subjects recruited as control and depressed subjects. A total of 84 subjects were recruited as of July 17th, 2023, excluding schizophrenia subjects. The Emory University Institutional Review Board and the Grady Research Oversight Committee granted approval for this study (IRB# 00105142). Interviewees were recruited from Research Match (researchmatch.org), a National Institutes of Health-funded online recruitment strategy designed to connect potential participants to research studies, and through Grady’s Behavioral Health Outpatient Clinic utilizing a database of interested research participants. Participants were aged 18 − 65 and were native English speakers. For the initial screening, interviewees were recruited for either a control group (no history of mental illness within the past 12 months) or a group currently experiencing depression. All diagnoses and group categorizations were verified and finalized by the overseeing psychiatrist and clinical team after the semi-structured interview.

Two subjects did not meet the inclusion criteria based on the information shared during the interview. Interviews from four subjects were accidentally interrupted or unrecorded due to technical issues with the subjects’ devices, and the recorded audio or video files from five subjects were corrupted or led to signal extraction errors in certain modalities (for example, rPPG extraction error due to large percentage of facial occupation due to large yaw angle). Hence, data from 73 subjects were included in the analyses. Table I shows the demographics of those included participants.

**TABLE I:** Demographics of the 73 subjects grouped by diagnoses. ± indicates the standard deviation of the measured variable. Subjects with current mental health conditions or a history of diagnosis within 12 months were grouped as “MHC”, while the rest were considered “Controls”. For gender, “M” refers to male, “F” refers to female, “NB” refers to non-binary, and “NA” refers to subjects who refused to answer. For race, “W” refers to White, “B” refers to Black, “A” refers to Asian, “H” refers to Hispanic, “O” refers to subjects who identify to more than one race, and “NA” refers to subjects who refused to answer. The year of education indicates the number of academic years a person completed in a formal program provided by elementary and secondary schools, universities, colleges, or other formal post-secondary institutions. High school completion usually corresponds to 12 years of education, whereas college completion usually corresponds to 16 years. *†* Education levels from two subjects were not recorded, and therefore the last entry is based on 22 Controls and 49 MHC individuals. No significant differences (Mann-Whitney, *p >* 0.05) were found in ages and years of education between Controls and MHC.

|  | Controls | MHC |
| --- | --- | --- |
| Number of Subjects | 22 | 51 |
| Age (Years) | $42.7 \pm 14.0$ | $36.6 \pm 13.2$ |
| Gender (M/F/NB/NA) | 9/13/0/0 | 10/38/2/1 |
| Race (W/B/A/H/O/NA) | 10/7/2/0/2/1 | 28/10/9/2/2/0 |
| Years of Education† | $17.3 \pm 4.6$ | $16.7 \pm 2.5$ |

### B. Interviews and measurements

The study team created the interview guide and protocol and have components that simulate a psychiatric intake interview [19]. All interviews were conducted remotely via Zoom’s secure, encrypted, HIPAA-compliant telehealth platform. Both Video and Audio were recorded. The remote interview was divided into three parts: 1) A semi-structured interview composed of a series of open-ended questions, a thematic apperception test [52], phonetic fluency test [53], and semantic fluency test [54], 2) a sociodemographic section, and 3) clinical assessments which included the Mini-International Neuropsychiatric Interview (MINI) 6.0 [55], McGill Quality of Life Questionnaire [56], General Anxiety Disorder-7 [16], and Patient Health Questionnaire-9 [17].

### C. Categorization

Subjects were categorized into four different two-class categorizations based on self-rated scales or clinicians’ diagnoses to evaluate feature performances in classifying categorizations generated from under different assessment procedures.

1. The first and primary categorization is control (n=22) vs. subjects with mental health conditions (MHC, n=51) based on diagnoses as shown in Table I. The latter included subjects diagnosed with any mental health condition currently or a history of diagnosis within 12 months, including disorders like MDD, major depressive episode (MDE), comorbid or primary GAD, PTSD, panic disorder, social anxiety, agoraphobia, psychotic disorder, manic illness, personality disorder, and obsessive-compulsive disorder. The control group included the remaining subjects, who could have mild suicidality, mild agoraphobia, mild substance abuse and dependence, or a remote history (not in the previous 12 months) of MDD/MDE and are not currently on an antidepressant medication. The following three categorizations only included a subset of subjects due to inclusion/exclusion criteria and missing self-rating results.
2. The second categorization is non-MDD-control (n=18) vs. MDD (n=38, past or current). Since both groups in the first categorization are heterogeneous, we used this categorization to assess further whether differences could be found between controls and subjects with past or current MDD. In this case, we defined non-MDD-control as subjects with no lifetime history of MDD or other mental health conditions (but could have mild suicidality, mild agoraphobia, mild substance abuse and dependence), while the MDD subjects have primary diagnoses of MDD but could include comorbid GAD, PTSD, panic disorder, social anxiety, agoraphobia, and substance use disorder.
3. The third categorization is moderately depressed (PHQ-9 scores *>* 10, n=24) vs. rest (PHQ-9 scores *<*= 10, n=43). PHQ-9 scores were not reported for six subjects, resulting in 67 subjects in this categorization. To evaluate performance in classifying the severity of self-rated depression symptoms, we used a PHQ-9 score-based categorization and adopted a cutoff of 10, which indicates moderate depression [17].
4. The fourth categorization is moderate GAD severity (GAD-7 scores *>* 10, n=16) vs. rest (GAD-7 scores *<*= 10, n=49). GAD-7 scores were not reported for 8 subjects, resulting in 65 subjects in this categorization. Similar to the third categorization, we used a GAD-7 score-based categorization and adopted a cutoff of 10, which indicates moderate anxiety and a reasonable cut for identifying cases of GAD [16], to evaluate performance in classifying the severity of self-rated anxiety symptoms.

## III. Methods

### A. Multimodal feature extraction

Figure 1 shows the proposed multimodal analysis framework that extracts visual, vocal, language, and rPPG time series signals at the frame or segment level, summarizes those time series with statistical and temporal dynamic features at the subject level (except for text embedding from the large language model, where the model directly generated subject-level embedding), and evaluates the performance of these features in clinical diagnoses or self-rated severity classification tasks described in section. II-C.

**Fig. 1:**
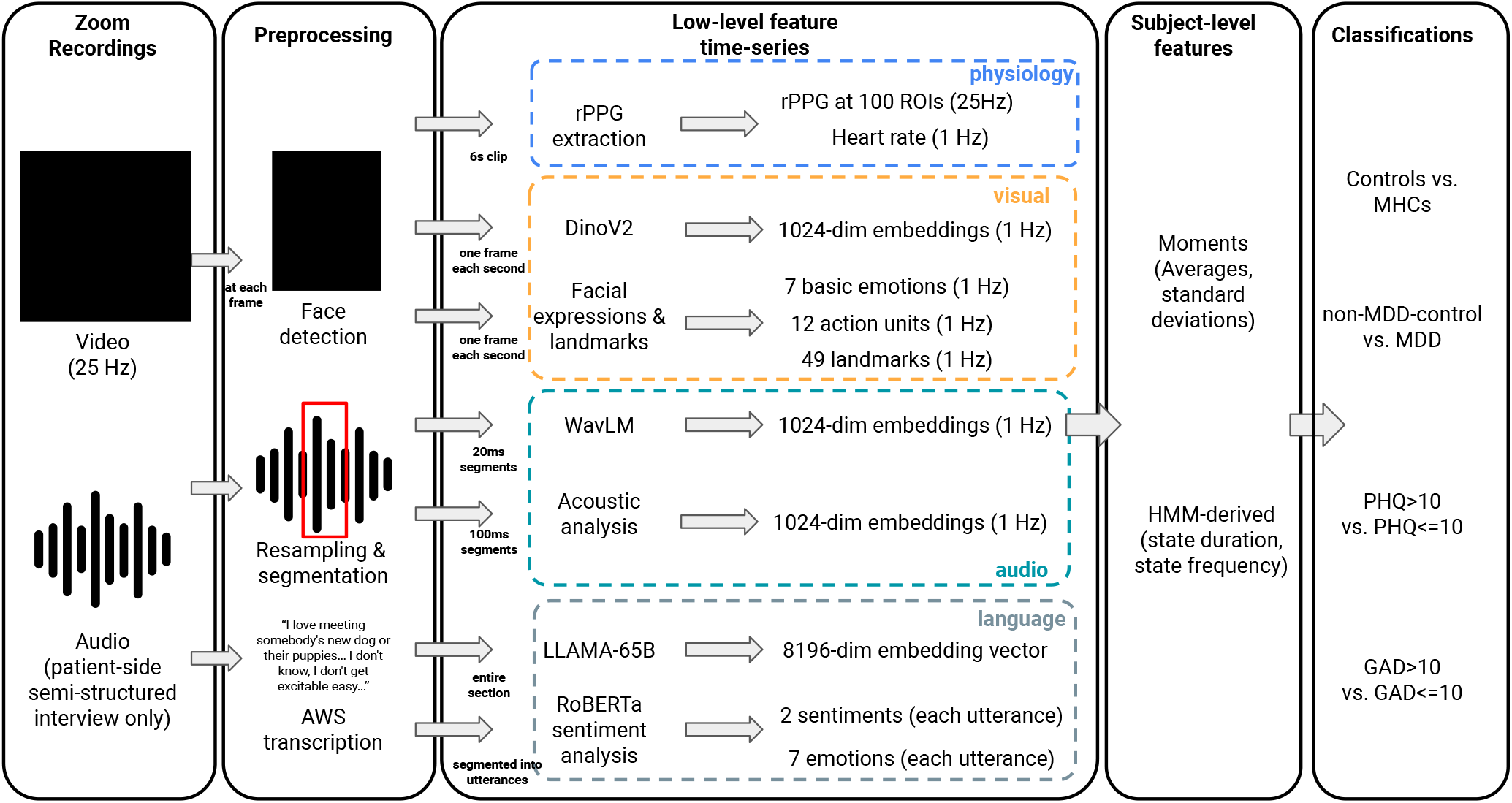
Overview of the processing pipeline. Colored dashed-boxes denote features from different modalities, including physiological, visual, audio, and language features.

#### 1) Facial expressions and visual patterns

We followed the CNN-based facial expression analysis framework we proposed in our previous work [20], [57]. For each frame of the recordings sample at 1 Hz (one frame per second), the face of the participant is detected with RetinaFace [58] using a ResNet-50 [59] backbone network trained on the “WIDER” face dataset [60]. The face detector achieved an accuracy of 95.5% on the “Easy” validation set in WIDER face dataset, where the faces were already much more difficult to detect than the faces in our use case. After segmentation, the face is fed into another CNN with VGG19 [61] structure, which was trained on the “AffectNet” dataset [62], to estimate the probabilities of the facial emotion expressed being into seven categories, namely neutral, happiness, sadness, surprise, fear, disgust, and anger. This facial emotion classifier was tested on the evaluation set in the AffectNet dataset and on a subset of the Radboud Faces Database (RaFD) [63]. It achieved an accuracy of 63.3% in the AffectNet evaluation set and 90.1% on the front-facing subset of the RaFD, while the accuracy of a random guess approach is 14.4%. and the agreement between two human annotators on the test set of AffectNet is only 60.7%.

To include facial behaviors less affected by cultural differences, we adopted JAA-Net [64] to recognize 49 facial landmarks and 12 facial action units [65] (AUs, or the individual components of facial muscle movement) expressed in the frame. JAA-Net is a deep learning model that combines CNN and adaptive attention module, and it achieved an average AU detection accuracy of 78.6% (including AU1, 2, 4, 6, 7, 10, 12, 14, 15, 17, 23, 24) and face alignment mean error of 3.8% inter-ocular distance on BP4D dataset [66] with threefold cross-validation.

In addition to manually-defined facial expression signals, including facial emotions, AUs, and facial landmark movements, a self-supervised large vision foundation model named “DINOv2” [67] was also used to extract general visual embedding of the segmented facial area. While video foundation models have better performance in short-video clips, the image foundation model was used because the average length of the video recorded in this study was significantly longer (one hour vs. a few seconds). DINOv2 is a vision transformer (ViT) [68] with one billion parameters trained on 1.2B unique images that achieved decent performance on video classification tasks with linear evaluation, including an accuracy of 90.5% on “UCF-101” dataset [69]. A 1024-dimensional visual embedding was generated from frames sampled at 1 Hz using the “ViT-L/14” [68] model.

#### 2) Language sentiments and representations

The patient-side audio files were transcribed into texts using Amazon Transcribe on HIPAA-compliant Amazon web services at Emory, following the protocol detailed in our previous study [70]. Similar to the audio analysis, only patient-side transcripts during the semi-structured interview section were used to avoid using subjects’ answers to sociodemographic or clinical assessment questions.

We have previously found different word use patterns in subjects with and without MDD using the linguistic inquiry and word count (version LIWC-22) dictionary [71]. Here large language models (LLMs) were used to identify the sentiments and extract general representations to better understand the subjects’ linguistic patterns. More specifically, three LLMs were used: (1) At the utterance level, a distilled RoBERTa model [72], [73] finetuned on 80% of 20k emotional texts (the rest 20% was used as the test set with an average accuracy of 66%) was used to recognize one of seven emotions including neutral, happiness, sadness, surprise, fear, disgust, and anger. (2) Also at the utterance level, another RoBERTa-based model finetuned on 15 diverse review datasets with a leave-one-dataset-out accuracy of 93.2% [74] was used to recognize positive or negative sentiment. Such fine-tuned utterance-level deep learning models have been found to generate effective representations in related contexts such as anxiety [75]. (3) LLAMA-65B [76], one of the state-of-the-art open-sourced decoder-only transformer models with 65 billion parameters which were trained on over one trillion tokens of texts, was used to generate an 8196-dimensional text embedding for the entire transcripts during the semi-structured interview.

#### 3) Vocal features and representations

Both manually defined acoustic features and general audio representations were extracted from audio files. Only patient-side audio during the semi-structured interview section was used to avoid the potential information leak directly from subjects’ answers to sociodemographic or clinical assessment questions in MINI or in self-rated questionnaires described in section II-B.

For manually defined features, *PyAudioAnalysis* [77] package was used to extract acoustic features at each 100ms window with 50% overlap, including zero crossing rate, energy, entropy of energy, spectral centroid/spread/entropy/flux/rolloff, Mel frequency cepstral coefficients, and 12 chroma vector and corresponding standard deviations. WavLM [78], which is a self-supervised audio foundation model with 316M parameters (“WavLM Large”) trained on 94k hours of audio, was used to extract general audio representations. It has shown state-of-the-art performance in the universal speech representation benchmark [79]. Recorded audio files were first resampled to 16k Hz and then segmented into non-overlapping 20ms segments following [78]. A 1024-dimensional audio embedding was generated for each 20ms segment using WavLM.

#### 4) Remote PPG cardiovascular features

Remote PPG signals were extracted from the video recordings using the *pyVHR* package [80], [81]. The facial skin areas were recognized in each frame using a CNN, 100 regions of interests (ROIs) were sampled, and the pixel values were averaged across the pixels in each ROI for each RGB channel, respectively. Then, an unsupervised method, named orthogonal matrix image transformation [82], was used to transform RGB values in one ROI to an estimated 25 Hz rPPG signal based on QR decomposition. The power spectral density of rPPGs at each ROI was computed in six seconds windows sliding every second, and the medians of the inverse of peak frequency (60*/*peak frequency) were used to estimate heart rates at every second.

Lastly, the averaged estimated rPPGs at each ROI were used to extract cardiovascular dynamic features using *PhysioNet Cardiovascular Signal Toolbox* [83] with a 300s window and a 30s sliding window. The cardiovascular dynamic features included time and frequency domain heart rate variability, acceleration and deceleration capacity, entropy measures, and heart rate turbulence measures. Highly tolerant rejecting thresholds were set to avoid rejecting high percentage of data, including setting lowest tolerable mean signal quality index (as defined in [83]) to be 0.1, allowing certain R-R intervals to be longer than ten seconds, allowing two neighboring R-R intervals to have a length difference of more than one second, and allowing a 30 seconds gap at the beginning of the PPG signals.

### B. Subject-level features and temporal analyses

Statistics of the time series extracted above were used as subject-level features. Both average and standard deviations over time were calculated for lower-dimensional (*<* 100) time series, including time series of facial expressions (facial emotions, AUs, and facial landmark locations sampled at 1 Hz), acoustic features (sampled at 20 Hz), language sentiments (sampled at each utterance), and estimated heart rates (sampled at 1 Hz). Only averages were calculated for higher-dimensional (*>* 100) time series, including time series of WavLM audio embedding and DINOv2 visual embedding. LLAMA-65B embedding of the entire semi-interviews was directly used as subject-level features.

In addition to nonparametric statistics, hidden Markov models (HMM) were used to model the dynamics of the low dimensional time series, and statistics (duration and frequency of inferred states) of the unsupervisely learned HMMs were used as subject-level features. An HMM with a Gaussian observation model and four states was learned for each modality separately using *SSM* package [84]. The number of states was selected because it represents the smallest number of states needed to model known different states: asymptomatic, symptomatic, uncertain, and padding states. Each time series of one modality from one subject *k, X*_*k*_, was considered as one noisy observation, where it is padded with zeros to the maximum temporal length *T*_*max*_ found from *X*_1_ to *X*_*N*_ (*N* = 73). i.e., *X*_*k*_ is a *T*_*k*_ × *d* with a feature dimension of *d* and a temporal length of *T*_*k*_ was padded (*T*_*max*_ − *T*_*k*_) × *d* zeros at the end, so all *X*_*k*_ has the same shape of *T*_*max*_ × *d*. The modality-specific HMM was then fitted on *X*, and the most likely hidden states *Z*_*k*_ with the shape of *T*_*max*_ × 4 were inferred for each sequence *X*_*k*_. Lastly, the time steps spent and the frequency (non-neighboring occurrences) of all four states were calculated for each subject and used as subject-level dynamic features.

### C. Classification analyses

We evaluated features generated from the above-described processes in four two-class classification tasks described in Section. II-C. Classification performances were measured by the average area under the receiver operating characteristic (AUROC) and the average accuracy in 100 repeated five-fold cross-validations, where subjects were randomly stratified into five approximately equally sized folds in each repetition.

#### 1) Demographic variables

Demographic variables, including one-hot-encoded race, one-hot-encoded gender, age, and years of education, were combined into a demographic feature vector for each subject and also evaluated as a benchmark in unimodal classification. However, demographic features were not considered in the multimodal classification.

#### 2) Unimodal evaluation

For each modality, statistics (averages and standard deviations) and HMM-derived features were evaluated separately using a logistic regression (LR) with *l*2 regularization or a gradient boosting decision tree (GBDT) classifier, depending on the dimensionality of the features, where LR was used for features with fewer than 100 dimensions.

#### 3) Multimodal fusion

Both early and late fusion of different modalities were considered. For early fusion, features from all modalities were concatenated into a single feature vector as the input to a GBDT classifier. For late fusion, the majority vote of each unimodal classifier was used as the multimodal classification results. To avoid noise from classifiers without classification power, we also compared the majority voting results from classifiers that showed non-random (defined as *AUROC >* 0.5) performance in the validation set (a 20% subset within the training fold). The non-random classifiers were re-trained with all the data in the training fold before being used for testing.

### D. Statistical Analyses

We used statistical tests to assess the differences in the probability distributions of features between different groups of subjects (such as groups described in Section. II-C and demographic groups) and the differences in performance resulting from different features. Mann-Whitney rank tests were applied between features or characteristics of different subject groups to determine whether significant differences exist between the two groups. McNemar’s test was used to evaluate the classification disagreement between pairs of classification settings. Wald Test was used to determine if a significant correlation was found between two variables. Significance was assumed at a level of *p <* 0.05 for all tests.

## IV. Results

### A. Unimodal feature patterns across groups

Here we performed a selected array of analyses of the clinically relevant patterns found in different modalities in different groups of subjects, providing additional objective evidence to previous clinical observations.

#### 1) Blunted visual affect and increased sadness in language

While “blunted affect” was mostly in the context of a negative symptom of schizophrenia, it has been widely reported in other mental disorders like MDD [85]–[87] and other non-psychotic disorders [88]. Measured by the sum of average AU intensities over the interview, we found that non-medicated subjects with current MDD had lower facial expressivity compared to non-MDD controls (Mann-Whitney, *p* = 0.04), and subjects with mental health conditions also had lower facial expressivity compared to controls (Mann-Whitney, *p* = 0.03). However, no differences in facial expressivity were found between subjects with past MDD and non-MDD controls, and no statistically significant linear correlations were found between facial expressivity and self-rated PHQ-9 or GAD-7 scores.

Through language sentiment analysis, neither was verbally blunted affect found in the MDD or MHC groups nor language expressivity correlate with self-rated scores. However, the average sadness level expressed in language was found to be higher in MDD groups compared to non-MDD-controls (Mann-Whitney, *p* = 0.02) and was positively correlated with PHQ-9 (Wald test, *ρ* = 0.31, *p* = 0.01) and GAD-7 (Wald test, *ρ* = 0.37, *p* = 0.002) scores. In comparison, the sadness level expressed visually did not increase in MDD groups.

#### 2) Increased acoustic spectral flux

The average spectral flux, defined as the squared difference between the normalized magnitudes of the spectra of the two successive frames averaged across the semi-structured interviews, was found to be positively correlated with PHQ-9 (Wald test, *ρ* = 0.26, *p* = 0.03) and GAD-7 (Wald test, *ρ* = 0.25, *p* = 0.04) scores, indicating a faster change of acoustic tones in subjects with more severe depression and anxiety symptoms.

#### 3) Increased complexity in heartbeat intervals

No significant alternation of average heart rate or standard deviation of heart rate during the interview was found between groups. The complexity of the heartbeat time series, measured by the area under the multiscale entropy curve, was significantly higher in non-medicated MDD groups compared to non-MDD-controls (Mann-Whitney, *p* = 0.01), consistent with previous findings using electrocardiogram [89], [90].

#### 4) Effect of medication

Compared to non-medicated MDD subjects, medicated MDD subjects showed a higher level of facial expressivity (Mann-Whitney, *p* = 0.05) and sadness (Mann-Whitney, *p* = 0.04), while only non-medicated subjects with current MDD showed a higher level of sadness through language compared to medicated subjects with current MDD (Mann-Whitney, *p* = 0.04). In addition, decreased heartbeat interval complexity (Mann-Whitney, *p* = 0.02) and increased standard deviation of heart rate (Mann-Whitney, *p* = 0.02) were observed with medication in subjects with past and current MDD compared to non-medicated MDD subjects, while the average heart rate remained similar between both groups.

### B. Dynamics inferred from HMM state duration and frequency

Dynamic features, including inferred HMM state duration and frequency, were found to be the most useful features in classification tasks, as shown in Table II, especially for facial expressions and rPPG modalities. Significant linear correlations were found between these dynamic features and PHQ-9/GAD-7 scores.

**TABLE II:** Classification performance of clinical diagnoses and self-rated depression/anxiety severity. Each column shows the performance of two-class classification using one of the four categorizations defined in Section. II-C in the same order. The average and the standard deviation of AUROCs and accuracies (in brackets) from a hundred randomly-split five-fold cross-validations are reported. The term *avgs* denotes averages, and *stds* denotes standard deviations. “random” indicates that the classifier performed no significantly better (McNemar’s test, *p >* 0.05) than random guessing (AUROC=0.5). The best classification performance in each task (column) achieved by a single modality was shown in bold text, while the second best was underlined. Multiple metrics were underlined or marked bold when no statistical significance (McNemar’s test, *p >* 0.05) between classifiers was found. The best classification performance in each task (column) achieved by multimodal fusion was shown in bold text. “†” indicates significantly better performance (McNemar’s test, *p <* 0.05) was achieved with the indicated feature type in this classification task (each column) compared to other unimodal features, where “‡” indicates significantly better performance (McNemar’s test, *p <* 0.05) was achieved with multimodal voting compared to using any unimodal features.

| Feature type | Metric | 1. Control vs. MHC | 2. Non-MDD-Control vs. MDD | 3. PHQ-9 > 10? | 4. GAD-7 > 10? |
| --- | --- | --- | --- | --- | --- |
| 1. Demographic variables | AUROC | 0.54 ± 0.04 | 0.54 ± 0.04 | 0.61 ± 0.03 | 0.57 ± 0.05 |
|  | Accuracy | 0.56 ± 0.04 | 0.57 ± 0.04 | 0.58 ± 0.04 | 0.60 ± 0.04 |
| 2. Facial emotions + AUs |  |  |  |  |  |
| 2.1 Avgs and stds | AUROC | random | random | 0.56 ± 0.06 | 0.55 ± 0.04 |
|  | Accuracy | random | random | 0.60 ± 0.05 | 0.62 ± 0.04 |
| 2.2 HMM features | AUROC | 0.65 ± 0.03 | 0.66 ± 0.04 | 0.61 ± 0.04 | **0.68 ± 0.05** |
|  | Accuracy | 0.64 ± 0.03 | 0.66 ± 0.04 | 0.60 ± 0.04 | **0.67 ± 0.03** |
| 3. DINOv2 avgs and stds | Both | random | random | random | random |
| 4. Language sentiment |  |  |  |  |  |
| 4.1 Avgs and stds | AUROC | 0.69 ± 0.03 | 0.66 ± 0.04 | 0.64 ± 0.05 | 0.63 ± 0.04 |
|  | Accuracy | 0.67 ± 0.04 | 0.68 ± 0.04 | 0.67 ± 0.05 | 0.64 ± 0.04 |
| 4.2 HMM features | AUROC | 0.62 ± 0.03 | 0.64 ± 0.04 | random | 0.65 ± 0.05 |
|  | Accuracy | 0.65 ± 0.03 | 0.60 ± 0.03 | random | 0.73 ± 0.04 |
| 5. LLAMA-65B | AUROC | 0.64 ± 0.07 | 0.53 ± 0.08 | 0.68 ± 0.04 | 0.64 ± 0.05 |
|  | Accuracy | 0.68 ± 0.05 | 0.59 ± 0.07 | 0.68 ± 0.03 | 0.70 ± 0.04 |
| 6. WavLM avgs and stds | AUROC | random | 0.58 ± 0.05 | 0.60 ± 0.06 | 0.59 ± 0.02 |
|  | Accuracy | random | 0.61 ± 0.05 | 0.64 ± 0.05 | 0.71 ± 0.02 |
| 7. Vocal acoustics |  |  |  |  |  |
| 7.1 Avgs and stds | AUROC | random | random | 0.68 ± 0.05 | random |
|  | Accuracy | random | random | 0.67 ± 0.05 | random |
| 7.2 HMM features | AUROC | 0.57 ± 0.05 | 0.51 ± 0.06 | 0.51 ± 0.05 | 0.53 ± 0.05 |
|  | Accuracy | 0.59 ± 0.04 | 0.53 ± 0.05 | 0.53 ± 0.05 | 0.60 ± 0.04 |
| 8. rPPG |  |  |  |  |  |
| 8.1 Cardiovascular features | AUROC | random | 0.55 ± 0.07 | 0.65 ± 0.04 | 0.56 ± 0.05 |
|  | Accuracy | random | 0.61 ± 0.05 | 0.60 ± 0.04 | 0.59 ± 0.04 |
| 8.2 HMM features | AUROC | **0.72 ± 0.05** † | **0.73 ± 0.05** † | **0.75 ± 0.03** † | **0.68 ± 0.04** |
|  | Accuracy | **0.76 ± 0.04** † | **0.73 ± 0.05** † | **0.71 ± 0.03** † | **0.67 ± 0.03** |
| 9. Fusion (row 2-8) |  |  |  |  |  |
| 9.1 Feature concatenation | AUROC | 0.63 ± 0.07 | random | 0.59 ± 0.05 | 0.53 ± 0.05 |
|  | Accuracy | 0.68 ± 0.05 | random | 0.61 ± 0.04 | 0.62 ± 0.04 |
| 9.2 Majority vote | AUROC | 0.70 ± 0.05 | 0.68 ± 0.07 | 0.75 ± 0.05 | 0.71 ± 0.05 |
|  | Accuracy | 0.73 ± 0.03 | 0.71 ± 0.04 | 0.71 ± 0.04 | 0.76 ± 0.03 |
| 9.3 Selected vote | AUROC | **0.82 ± 0.04** ‡ | **0.77 ± 0.02** ‡ | **0.82 ± 0.04** ‡ | **0.72 ± 0.04** ‡ |
|  | Accuracy | **0.75 ± 0.03** ‡ | **0.76 ± 0.01** ‡ | **0.74 ± 0.04** ‡ | **0.75 ± 0.03** ‡ |

Figure 2 shows the correlation plots between the frequency of the states in emotion and heart rate time series. The padding states (described in section III-B) from rPPG and facial expression HMMs were omitted as they would only present once (frequency=1) as the padding in the end. Statistically significant positive correlations were found between all non-padding state frequency and self-rated scores except emotion state 2, indicating a higher switching rate between hidden states may be related to more severe depression and anxiety symptoms.

**Fig. 2:**
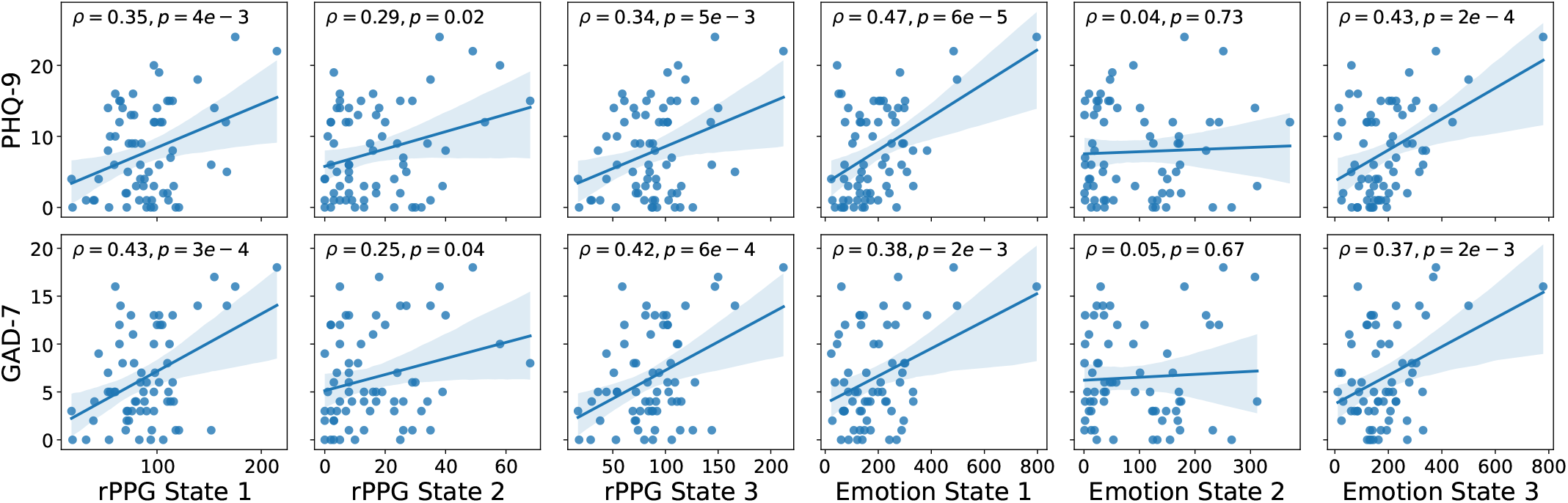
PHQ-9 and GAD-7 scores vs. rPPG and facial expression HMM state frequencies. Each subfigure shows the scatter plot between self-rated scores and the frequency of a learned HMM state, along with a linear regression model fit with the 95% confidence interval. The top row shows how those learned state correlate with PHQ-9 scores and the bottom row shows how they correlate with GAD-7 scores. Texts in each subfigure denote the Pearson correlation coefficient (*ρ*) and the p-value using the Wald test.

### C. Classification performance

Table II shows the classification performance of both clinically diagnosed and self-rated mental health disorders using static and dynamic features from vision, audio, language, and physiology. Each column shows the performance of two-class classification using one of the four categorizations defined in Section. II-C in the same order. The best-performing features achieved an AUROC of 0.68 to 0.75 in unimodal classification tasks, while the selected majority voting described in Section III-C3 achieved an AUROC of 0.82 in detecting current or recent (last 12-month) mental disorders, an AUROC of 0.77 in detecting past and current MDD, an AUROC of 0.82 in detecting PHQ-9 based moderate depression, and an AUROC of 0.72 in detecting GAD-7 based moderate anxiety disorder. Late fusion using selected majority voting (row “9.3”) outperformed early fusion with the direct concatenation of features (McNemar’s test, *p* ≪ 0.01) due to the extremely high dimensionality of the concatenated features.

While demographic variables achieved higher than random performance in all four tasks, we found they were not strong predictors of mental health disorders compared to the proposed features, as shown in row “1” in Table II.

#### 1) Moments and dynamics of facial expressions revealed mental states but general visual patterns did not

While we also extracted facial landmarks as described in Section III-A1, we found adding static statistics of facial landmarks or including facial landmarks in the HMM modeling deteriorated the performance. Row “2” in Table II shows the performance using just the statistics of facial emotions and AUs. Interestingly, the average and standard deviation of facial expressions failed to classify clinical diagnoses but successes in classifying self-rated depression and anxiety. In comparison, the temporal properties derived from HMM resulted in significantly better (McNemar’s test, *p* ≪ 0.01) classification performance, except for self-rated depression detection. Lastly, using the temporal dynamics of facial expressions achieved the best performance in self-rated anxiety in all modalities.

In comparison, visual embedding generated from DINOv2 failed to generalize to this specialized dataset and did not achieve non-random classification in any of the tasks.

#### 2) Language sentiments beat general language representation in small and specialized dataset

Compared to other modalities, language features were extracted at a lower sampling rate (at each utterance or the entire semi-structured interview), while LLMs were able to abstract the texts into much shorter sequences of features or even into a single vector when LLAMA-65B was used. The average and standard deviation of the language sentiments achieved the best performances compared to static features of other modalities. Using HMM to model the sentiment dynamics did not improve performance, as shown in all other modalities (comparing rows “4.2” and “4.1”). These results showed that part of the dynamics expressed through the words was already captured by LLM and abstracted into utterance sentiments, and the sentiment dynamics over multiple utterances might not be as important.

Additionally, while using LLAMA-65B embedding showed decent performance compared to other non-language modalities, using language sentiments achieved similar or better results in all tasks. This showed that general language representation might not be as useful as disorder-related sentiment analysis, especially in smaller and highly-specialized datasets, as demonstrated in this study, and suggested in related work on text-based depression and personality detection [91], [92].

#### 3) Vocal features were under-performing compared to other modalities

While many previous studies [26], [27], [93] have shown that vocal features are useful in detecting depression and anxiety disorder, in this study, other modalities outperformed both spectral/entropy-based acoustic features and general speech representation from WavLM except in self-rated depression detection.

#### 4) HMM modeled dynamics were more informative compared to cardiovascular features for highly noisy rPPG signals

As shown in row “8.1” in Table II, using cardiovascular features yielded inferior performance compared to other modalities. The key reason is the estimated rPPG signals were highly noisy at each ROI or after averaging across all ROIs, which led to errors (such as peak detection error) in downstream cardiovascular feature calculations. On average, 25.8% of the estimated rPPGs were not used for downstream analyses even with highly tolerant rejecting thresholds as described in Section III-A4. Using HMM-derived features from modeling heart rate time series resulted in the best or second-best performance in all four tasks among all unimodal approaches, reaching AUROCs from 0.68 to 0.75.

## V. Discussion and conclusion

In this work, we performed a thorough multimodal analysis on 73 subjects using remotely-recorded telehealth interviews and showed that the facial, vocal, linguistic, and cardiovascular features extracted from these audiovisual recordings could reveal informative characteristics of both clinically diagnosed and self-rated mental health status. The results provided early evidence of the usefulness of multimodal digital biomarkers extracted from low-cost and non-lab-controlled data with minimal hardware limitations. Comparisons were made between different modalities and between features derived from the latest transformer-based foundation models and more defined features derived from traditional methods, offering insights on which modalities and methods might be most suited for automated remote mental health assessments.

### A. Performance of different modalities

When comparing the classification performance using features extracted from different modalities, it is clear that overall physiological characteristics outperformed other manually-defined or data-driven behavioral characteristics. Heart rate dynamics were highly relevant in classifying self-rated and clinician-diagnosed disorders, even though the heart rates were estimated indirectly from light changes on the face. While it is not surprising to find associations between cardiovascular dynamics and psychiatric disorders, as shown in previous studies of neurobiological mechanisms [94] and statistical analyses [95], [96], the results raised questions on the behavioral features extracted in this study. More investigations are needed to answer whether they underperformed because the current state-of-the-art models cannot capture enough information in remote interviews, or behavioral signals are not as useful as physiological signals in telehealth settings, even for human experts.

Among behavioral modalities, overall facial and language patterns led to better classification performance than patterns derived from audio, although the latter resulted in a comparable performance in detecting self-rated depression. While overall facial and language patterns led to similar levels of performance, it is worth noting that they performed very differently in different tasks, suggesting the same modality might perform differently for different mental health assessment tasks. For example, facial expression dynamics were much more useful in detecting self-rated anxiety than self-rated depression, yet similarly useful in detecting clinical MDD. On the contrary, Language embedding was more powerful in detecting self-rated disorders than clinically diagnosed disorders. These findings caution us on translating and interpreting results found using self-rated or self-reported scales directly for clinical applications, where the categorization criteria and process are different, in addition to the subject distribution shift, which was not shown in this study (as they were evaluated on the same group of subjects).

### B. The use of foundation models

Foundation models have gained enormous popularity in the last few years with the rapid development of pre-training and self(semi)-supervised training methods [97], [98], especially since the release of OpenAI’s ChatGPT. LLMs, along with visual [67], audio [78], and multimodal [99] foundation models were widely applied in many disciplines, including the mental health domain, but primarily limited to language analyses and self-rated (self-reported) conditions [100], [101]. By comparing the direct use of unimodal foundation model-generated embedding to manually defined features from the same modalities, we showed how they perform in more clinically-relevant tasks under the telehealth settings.

The statistics of the visual embedding from DinoV2 were not at all useful in detecting mental health disorders. This finding was partially expected because the majority of extracted general visual representations would be more relevant to the texture and appearance of the face, especially after averaging, while the dynamics of the high-dimensional embedding would be hard to find with a limited number of recordings (discussed in more details in section V-C below). Preliminary results using other vision foundation models in this dataset did not show better classification performances either, including using models tuned for facial representation (“FaRL” [102]) and for facial video representations (“MARLIN” [103]).

Although audio embedding from WavLM only outperformed acoustic features slightly here, it showed the potential of using the general audio embedding from a more diversely pre-trained audio foundation model in datasets with more subjects. Interestingly, general text embedding of the entire semi-structured interview from LLAMA-65B performed similarly when compared to sentiment analyses considering the extremely high feature dimensionality and the small number of recordings. With the rapid development of LLMs and the inclusion of more diverse training texts, such as the recent release of LLAMA2 [104], general LLMs could potentially outperform fine-tuned task-specific LLMs in mental health assessment tasks in the near future.

### C. Limitations and future directions

Several limitations of this study need to be acknowledged, as they provide valuable insights into the boundaries of our findings and potential directions for future studies.

First, the number of subjects (n=73) and their heterogeneity might limit our findings’ generalizability. While the number of subjects will grow as we keep collecting data following our previous protocol [19], the heterogeneity issue might not be easily addressed. Although we recruited subjects with clear inclusion/exclusion criteria and further excluded subjects after the interview if they did not fall into our criteria, the intrinsic nature of high comorbidity levels in different mental disorders makes it difficult to recruit a “clean” cohort of subjects with clear diagnoses of a single type of disorder. Another heterogeneity came from medication status, which has been known to affect both behaviors and physiology of the patients [105]–[107]. That said, we believe the heterogeneity could be partially addressed as the number of subjects grows because analyses of smaller and more well-defined groups, which do not have enough samples currently, could be performed. As the number of subjects grows, models used for feature extractions could be potentially fine-tuned on the targeted population instead of being only trained on open-access datasets, which could further close the gap in getting accurate features from the target population.

Second, potential bias in the feature extraction process might exist. The facial expression model used in this study was evaluated in our previous research [57], but the features from other modalities were extracted using open-access models that may bias towards certain demographic groups, leading to potential skew in the findings. For example, LLAMA is reported to be biased in religion, age, gender, and other aspects as it was trained with internet-crawled data [76]. A thorough bias analysis must be performed in a future study before applying it clinically.

Third, the unimodal and multimodal classification and fusion methods used in this study could be improved given a larger and more densely labeled dataset. Only one label (per task) was available for the entire recording, which made it difficult to apply temporal models such as recurrent neural networks or transformers to directly classify high-dimensional time series with thousands to tens of thousands of steps. A potential solution is augmenting the labels by segmenting the recordings into shorter clips and assigning the same labels for all the clips from the same subjects. However, this process may lead to many false positives as the symptoms or cues relevant to mental health might only appear a few times. Similarly, a multimodal transformer could be potentially used for fusion, provided the label sparsity challenge is addressed. A potential future direction is to label the entire recording more frequently in time. For example, simple measurements like self-rated or clinician-rated levels of distress could be adopted. Another potential direction is to utilize the potential improvement in pre-trained foundation models. For instance, LLMs with larger context windows might enable few-shot classification by including a few examples of transcripts and categorizations in the prompt.

### D. Potential clinical applications

With larger and more diverse samples, we see considerable potential clinical utility for the proposed multimodal objective assessment approach (and future applications informed by this technology) in several areas: 1) deepening understanding of psychopathology and outward manifestations of symptoms, 2) utility for diagnostic purposes, 3) assessing changes in symptoms longitudinally for the same patient, and 4) for patient self-report, engagement, and empowerment. First, this technology has the potential to better objectify and quantify core signs and symptoms of certain mental health conditions, like affective flattening or tangential speech. Second, this technology has the potential to augment the initial diagnostic process for clinicians in both research and clinical settings. Developing real-time reporting of digital biomarker outputs in the form of a dashboard may help clinicians may hone into a certain line of clinical questions to better help establish a diagnosis. This technology may have a role in reducing bias and discrimination in the diagnostic process, as currently, the preponderance of evidence suggests that Black/African American individuals and Hispanic individuals are disproportionally diagnosed with psychotic disorders [108]. In time, combining digital biomarkers in addition to other blood-based and imaging markers, could play a potential role in subtyping mental health conditions according to treatment response or identifying individuals at risk who might develop the condition [109]. Third, applications of this technology can help clinicians and researchers to assess changes in symptoms over time for the same patient. This is crucial for the health care team to understand if the treatment plan is working and may help to accelerate measurement-based care efforts and overcome some of the barriers to implementation [110]. Additionally, accurate assessment is the cornerstone of clinical research studies which ultimately determines whether new treatments are approved, and unreliable assessments can have significant consequences to the study and to the field more broadly [111]. Finally, future applications informed by this technology can play an important role in empowering patients to participate in self-assessment and ongoing monitoring of their symptoms. Such applications may help to improve the accessibility/timeliness of assessments and even reduce stigma around mental health [112].

## Data Availability

Data cannot be shared publicly as it contains personal health information such as diagnoses and demographics and personal identified information such as video recordings of the participants' faces. They cannot be used or shared beyond the scope of this study due to the protection of patient confidentiality and the ethical restrictions imposed by Emory Institutional Review Board. Data are available from the Emory IRB (contact via) for researchers who meet the criteria for access to confidential data.

